# Clinical benefits of modifying the evening light environment in an acute psychiatric unit: A single-centre, two-arm, parallel-group, pragmatic effectiveness randomised controlled trial

**DOI:** 10.1101/2024.03.21.24304657

**Authors:** Håvard Kallestad, Knut Langsrud, Melanie Rae Simpson, Cecilie Lund Vestergaard, Daniel Vethe, Kaia Kjørstad, Patrick Faaland, Stian Lydersen, Gunnar Morken, Ingvild Ulsaker-Janke, Simen Berg Saksvik, Jan Scott

**Affiliations:** Department of Mental Health, Norwegian University of Science and Technology, Trondheim, Norway; Division of Mental Health Care, St. Olavs hospital, Trondheim University Hospital, Trondheim, Norway; Department of Health Promotion, Norwegian Institute of Public Health, Bergen, Norway; Institute of Neuroscience, Newcastle University, UK

## Abstract

**Background:** The impact of light exposure on mental health is increasingly recognized. Modifying inpatient evening light exposure may be a low-intensity intervention for mental disorders, but few randomized controlled trials (RCTs) exist. We report a large-scale pragmatic effectiveness RCT exploring whether individuals with acute psychiatric illnesses experience additional benefits from admission to an inpatient ward where changes in the evening light exposure are integrated into the therapeutic environment.

**Methods and findings:** All adults admitted for acute inpatient psychiatric care over eight months were randomly allocated to a ward with a blue-depleted evening light environment or a ward with standard light environment. Baseline and outcome data from individuals who provided deferred informed consent were used to analyze the primary outcome measure (differences in duration of hospitalization) and secondary measures (differences in key clinical outcomes). The Intent to Treat sample comprised 476 individuals (mean age 37; 41% were male). There were no differences in the mean duration of hospitalization (6.7 vs. 7.1 days). Inpatients exposed to the blue-depleted evening light showed higher improvement during admission (Clinical Global Impressions scale-Improvement: 0.28, 95% CI: 0.02 to 0.54; p=0.035, Number Needed to Treat for clinically meaningful improvement (NNT): 12); lower illness severity at discharge (Clinical Global Impressions Scale-Severity: –0.18, 95% CI: –0.34 to – 0.02; p=0.029, NNT for mild severity at discharge: 7); and lower levels of aggressive behaviour (Broset Violence Checklist difference in predicted serious events per 100 days: –2.98; 95% CI: –4.98 to –0.99; p=0.003, NNT: 9). Incidents of harm to self or others, side effects, and patient satisfaction did not differ between the lighting conditions.

**Conclusions:** Modifying the evening light environment in acute psychiatric hospitals according to chronobiological principles does not change duration of hospitalizations, but can have clinically significant benefits without increasing side effects, reducing patient satisfaction or requiring additional clinical staff.

---

Light is the most critical environmental factor for circadian rhythmicity and research over several decades indicates that manipulating light and dark exposure may improve clinical outcomes.^1–3^ The study of chronotherapies has been galvanised by new research demonstrating that the circadian effect of light on humans is primarily mediated by intrinsically photoreceptive retinal ganglion cells (ipRGC) that have peak sensitivity to blue light.^4–7^ This discovery indicates that it may be feasible to achieve clinical improvements by specifically blocking the blue part of the light spectrum in the evening, so-called virtual darkness, without recourse to prolonged sensory deprivation which is employed in dark therapies.^8^ For example, several studies, including small-scale randomised controlled trials (RCTs), have examined the potential benefits of the adjunctive evening use of ‘blue blocking glasses’ (BBG) compared with standard treatment for individuals with insomnia, delayed sleep phase, major depression, post-partum disorders, bipolar disorder, and mania.^1,9–15^ Overall, findings are inconsistent. However, it is unclear whether this is due to a lack of clinically significant effects in certain subgroups, study methodology, e.g., sample size or case mix, or the specific outcome measures employed to define benefit. For example, many trials focused only on mood and/or sleep disorders and affective disorders, but it is possible that BBG might influence other acute psychiatric symptoms, such as agitation and aggression or suicidality, across a broad range of mental disorders.^1,9–15^ Alternatively, the effectiveness of BBG may be attenuated because many individuals, especially acutely mentally ill inpatients, struggle to adhere with the interventions or lack capacity to participate in research.

One way to increase access to chronotherapy for severe mental disorders would be to follow the example of some new-build general hospitals and design psychiatric inpatient facilities with dynamic ‘circadian’ lighting systems that change light spectrum and intensity according to time of day, e.g., evening blue depleted light.^16^ A particular appeal of such innovations is that, once installed, the intervention is ‘low intensity’ in as much as no additional clinical staff are required to manage the system. Our own research has shown that changing the evening light environment in an acute psychiatric unit has positive effects on the circadian, sleep and neurocognitive systems of healthy young adult volunteers.^17,18^ However, it is unknown whether any change would translate into better clinical outcomes. Several mental health inpatient projects are underway, but there is limited research on the potential benefits of exposing trans-diagnostic inpatient populations to dynamic, programmable lighting conditions. To our knowledge, only two small-scale RCTs have been published.^19,20^ Canazei and colleagues (2022) used actigraphy to monitor sleep and rest-activity rhythms in 30 individuals with depressive disorders admitted to inpatient rooms with circadian or standard lighting conditions. The study showed that although exposure to the experimental lighting was associated with some significant improvements in selected sleep and circadian metrics, the duration of admission did not differ between groups (20 *vs*. 21 days). Okkels and colleagues (2020) randomly allocated 54 individuals admitted to an affective disorder unit to a pre-set circadian lighting condition where light intensity and spectrum is changed throughout the 24h cycle or standard lighting environment. Most inpatients had an ICD-10 diagnosis of depression, but a minority had mania, anxiety, or personality disorders. The intervention group demonstrated non-significantly greater improvements in sleep quality and other clinical symptoms, and no differences in length of stay (22 *vs* 19 days in the control group).

In summary, uncertainties exist regarding the potential extent or magnitude of any improvements in clinical outcomes, including duration of general psychiatric acute admissions, that may be associated with exposure to evening blue depleted light alongside usual inpatient care and treatment. The only consistent published finding is the lack of clinically significant side effects or adverse events.

However, the small sample sizes and selective recruitment strategies mean that many inpatient sub-populations were under-represented (e.g., psychotic disorders).^21^. Lastly, the trials of acutely ill patients employed eligibility criteria that *de facto* limited the generalizability of findings to real world settings, such as the permanent exclusion of inpatients with severe symptoms, complex presentations, or impaired capacity to give immediate consent.

## Aims

Given the above, we employed an ethically approved, deferred consent procedure that permitted acute admissions to be randomized to adult inpatient care in a ward with programmable dynamic lighting with a blue-depleted evening light environment (Blue-depleted LE; experimental group) or a similar ward with standard light environment (Standard LE; control group). This article reports the key findings, namely:

*a*) *Primary outcome:* any differences in the mean duration of admission in days per individual according to group allocation (i.e., evening light environment).
*b*) *Secondary outcomes:* any between-group differences in clinical symptom and function ratings, risk of or actual incidents of harm to self or others, patient satisfaction and side effects.

## Methods

The reporting of this single-centre, two-arm, parallel-group, pragmatic effectiveness randomised controlled trial follows the CONSORT guidelines.^22^ The trial was an investigator-initiated study sponsored by St. Olavs hospital, Trondheim University hospital, Trondheim, Norway.

Here, we give an overview of the trial methodology whilst the online Supplementary Information provide other relevant information including the CONSORT checklist and further details of consent procedures, data management, and analyses.

### Ethics statement and registration

The trial was prospectively registered to the Regional Committee for Medical and Health Research Ethics in Central Norway, May 8^th^ 2018 (REK 2018/946). The trial was approved on June 6^th^ 2018 and all participants gave a written informed consent. Upon approval by the Committee, the trial registration and study protocol were made publicly available in a searchable database (see Supplementary Information, p. 62). The RCT was listed on the Current Research Information System in Norway July 19th, 2018 (CRISTIN ID 602154). The initial trial registration and protocol (version 1) is available in the Supplementary Information, p. 3. An updated protocol was submitted to the Committee on Oct 17^th^ 2018 and was later registered on clinicaltrials.org and published (version 2, see supplementary information p. 25). Due to logistical issues in the study group, and the need to start inclusion based on seasonal variations in daylight, the trial was retrospectively registered on clinicaltrials.org on Dec 28^th^, 2018 (Clinicaltrials.gov NCT03788993; see Supplementary Information, p. 62 for details) which was updated with a detailed Statistical Analyses Plan (SAP) that was written and published prior to unblinding (see Supplementary Information, p. 40). No major changes were made to the study design between the prospectively registered protocol and the current manuscript.

### Study design and participants

The sample comprised individuals whose acute clinical presentation required inpatient care in the newly built 40-bedded adult psychiatric unit at St. Olavs Hospital, Trondheim, Norway (catchment area 300,000). The target was to recruit a minimum of 400 adults from the acute psychiatric admissions that occurred at any time and on any day of the week from October to March (to minimize the effect of seasonal variation in daylight; see Supplementary Information, p. 62). There were two recruitment periods: October 23^rd^, 2018, to March 29^th^, 2019; and October 1^st^ to November15^th^, 2019 (see Supplementary information, 63).

An independent randomization procedure was developed and managed by the Unit for Clinical Research at the Faculty of Medicine and Health Sciences, NTNU. As soon as the decision to admit an individual was confirmed, the person was randomly allocated to one of the two arms of the RCT on a 1:1 basis via a web-based program using blocks of randomly varying size. Randomization could be instigated at any time of the day or night without consultation with the hospital ward staff (regarding bed availability, case-mix, or staffing levels, etc.) and hospital staff could not influence the allocation procedure in any way.

### Eligibility criteria (pre– and post-randomization)

To minimize exclusions, we employed a post-randomization, deferred (delayed) consent procedure as utilized in many RCTs aiming to recruit representative samples of severely ill patient populations (also see Supplementary Information, p. 37).^23–25^

With ethical approval, the RCT eligibility criteria were as follows:

Inclusion criteria: All individuals aged >= 18 years who were admitted to the acute inpatient unit during the recruitment period were eligible for randomization. Individuals who were re-admitted were also eligible for re-randomization.

Exclusion criteria: There were no pre-randomization exclusion criteria, but individuals could be withdrawn from the study post-randomization.

Immediately post-randomization, there were two options for withdrawing a participant from the RCT:

1) Lack of availability of rooms in the ward to which the individual was allocated, i.e., the randomization process could not be completed.
2) Clinical imperative: a senior clinician decided that it was inappropriate to admit an individual to the room to which they are randomized. Reasons for which could be because admission to the allocated room might adversely affect the clinical case mix within the ward or compromise patient safety (see Supplementary Information, p. 64 for examples).

During the admission, or at the point of discharge, withdrawal from the RCT could occur because:

1) Lack of consent. The individual was unwilling to give written informed consent during an admission, according to the deferred consent procedure; was unable to give informed consent for the duration of the study, i.e., they persistently lacked mental capacity; or the consent procedure was incomplete: the individual had been discharged early or had an unplanned discharge so they were not approached about participation or had given verbal, but not written consent.
2) A patient could be withdrawn from the study if they were absent for >24 hours from the ward to which they were randomized. This could be in instances where patients were transferred to a somatic hospital ward for several days; or clinicians instigated transfer to another psychiatric ward, etc.
3) An individual could decline to participate or withdraw their consent at any stage of the study and/or a mental health professional could recommend withdrawal of an inpatient from the RCT if they had any clinical concerns regarding an individuals’ participation. Potential reasons could be if a clinician believed a patient had experienced an RCT-related adverse event.

### Experimental and control conditions

The construction of the 40-bedded psychiatric inpatient unit was initiated in 2015 and finished in December 2017. The unit is divided between two wards built around two atriums (see Supplementary Information, Fig. 1S). Each hospital ward has the same staffing levels, layout, and facilities and five rooms in each ward are designated as ‘psychiatric intensive care’ beds. The staff rotated between the wards every six weeks.

**Fig. 1:**
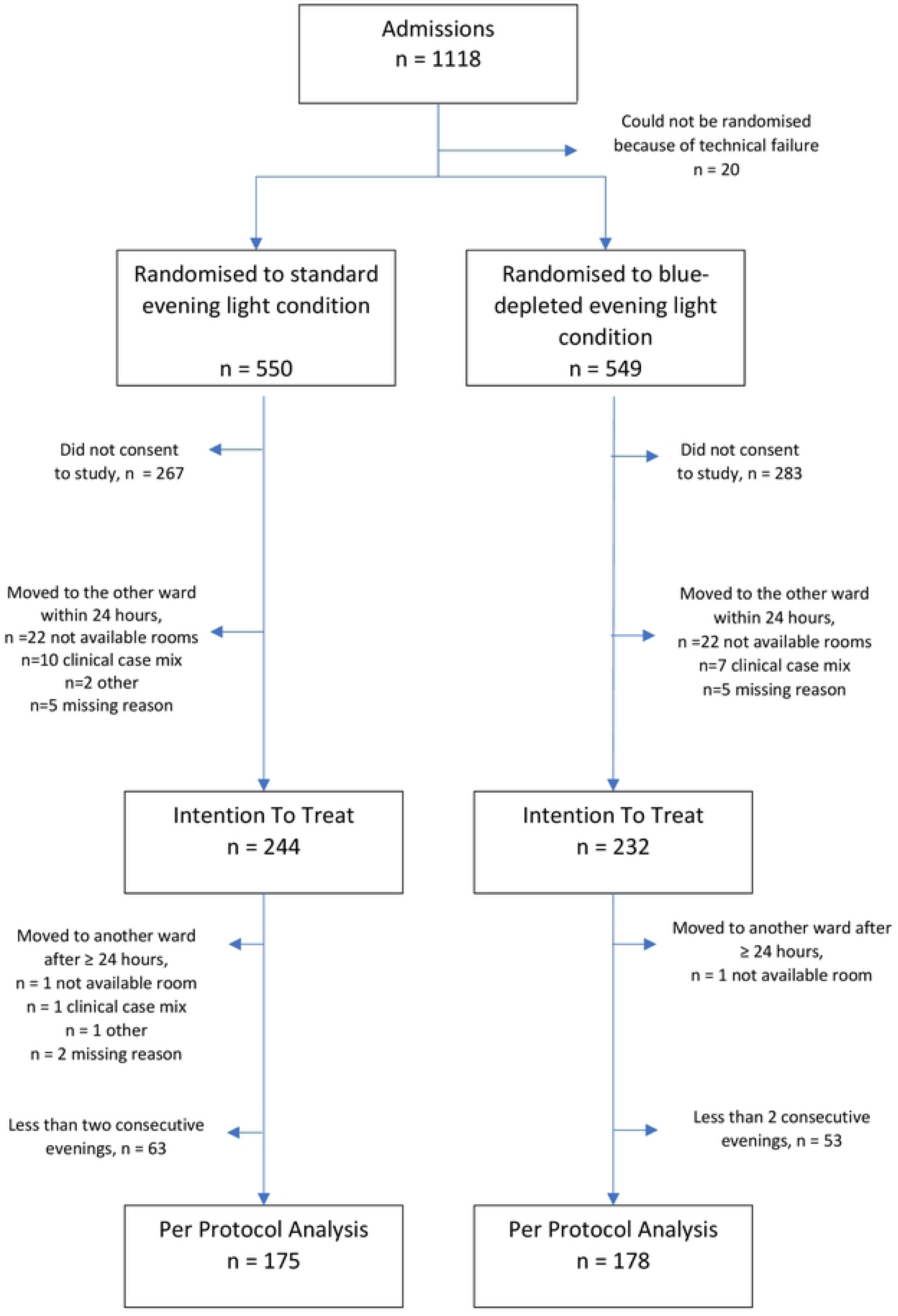
CONSORT flow diagram

The lighting fittings and fixtures were identical in both wards (Glamox AS, Oslo, Norway), while the diodes were different. During the RCT, the light intensity (photopic lux) was similar across the inpatient unit, but individuals were exposed to a different spectrum of evening light in each ward. See Vethe et al.^18^ for details about the composition of the evening LE in the two wards. We confirmed that LE was similar to Vethe et al.^18^ before the recruitment was initiated.

a) Experimental condition (Blue-depleted Evening Light Environment): the ward had tunable light emitting diode (LED) fixtures and the amount of blue light in the ward was tested prior to commencing the RCT. At 18:00 hours (h) the lighting underwent a 30-minute transition during which the green and blue LEDs were dimmed to produce blue-depleted amber coloured lighting. At 06:50h a 10-minute transition programme changed the light colour to normal indoor lighting (3000 Kelvins of colour temperature) which then continued until 18:00h. The light intensity was dimmed to 20% (of the maximum) from 23:00h to 6:50h. As well as the LED system, blue-blocking window filters were deployed in the evening. All television sets had permanent blue-blocking filters and the outdoor area had external lights that block blue light. Use of electronic media was not restricted (unless an individual treatment plan limits access), but patients were provided with blue-blocking screens that could be attached to the front of all electronic devices (lowbluelights.com). If a patient left the blue-depleted unit after 18:30 they were offered blue-blocking glasses to wear (circadianeyewear.com).
b) Control condition (Standard Light Environment): the ward had normal indoor hospital lighting installed. The light intensity was dimmed to 20% (of the maximum) during the night (from 23:00h to 06:50h).

### Assessments

Full details of all assessments employed in the main and ancillary studies are reported in the published protocol.^1^ Here, we briefly describe the assessments employed in the RCT.

#### Diagnosis

A preliminary diagnosis/diagnoses was recorded at admission, but the analyses used the consensus discharge diagnosis/diagnoses of mental disorders according to the ICD-10 ‘criteria for research’ (Chapter F) (World Health Organization; WHO).^26^

#### Baseline assessment

At intake, the following information was recorded:

a) age, sex, ethnicity, marital status, living situation, years of education, employment status.
b) other key characteristics include type of admission (voluntary or involuntary), current alcohol and substance use, risk of or actual harm to self or others, physical health status, disrupted sleep (operationalized as disturbances >=3 nights per week the last 30 days before admission), and medications used before/at admission (categorized according to the WHO class of medication; Supplementary Information, Table 1S).
c) details of past psychiatric history, specifically recording total number of psychiatric admissions and number of inpatient bed-days in the two years prior to the index admission, and forensic history (including history of violence).

**Table 1.** Summary of participant demographics and clinical characteristics for patients in the Intention to Treat population, randomized to admission in a standard light environment or evening blue-depleted light environment.

| Variable | Blue-depleted evening light environment (n=232) |  | Standard light environment (n=244) |  | Total (N=476) |  |
| --- | --- | --- | --- | --- | --- | --- |
| *Demographic information* |  |  |  |  |  |  |
| Male, n (%) | 102 | (44.0) | 91 | (37.3) | 193 | (40.5) |
| Age, years, mean (SD) | 37.1 | (13.3) | 37.2 | (14.4) | 37.2 | (13.9) |
| Ethnicity, European, n (%) | 189 | (81.5) | 180 | (73.8) | 369 | (77.5) |
| Education, primary school only, n (%) | 44 | (19.0) | 49 | (20.1) | 93 | (19.6) |
| Employment status, full-time, n (%) | 39 | (16.8) | 52 | (21.3) | 91 | (19.1) |
| Living situation, no fixed abode, n (%) | 17 | (7.3) | 15 | (6.1) | 32 | (6.7) |
| *Diagnostic data for ICD-10 mental disorder diagnoses, criteria for research* |  |  |  |  |  |  |
| Number of diagnoses (n, %) |  |  |  |  |  |  |
| 0 | 21 | (9.1) | 12 | (4.9) | 33 | (6.9) |
| 1 | 123 | (53.0) | 145 | (59.7) | 268 | (56.4) |
| 2 | 65 | (28.0) | 64 | (26.4) | 129 | (27.2) |
| 3+ | 23 | (9.9) | 22 | (9.0) | 45 | (9.5) |
| Primary diagnoses, according to ICD-10 chapters, n (%) |  |  |  |  |  |  |
| Affective disorders | 60 | (25.9) | 61 | (25.1) | 121 | (25.2) |
| Psychotic disorders | 36 | (15.5) | 51 | (20.9) | 87 | (18.3) |
| Personality disorders | 23 | (9.9) | 32 | (13.2) | 55 | (11.6) |
| Addiction disorders | 41 | (17.7) | 29 | (11.9) | 70 | (14.7) |
| Anxiety disorders | 24 | (10.3) | 29 | (11.9) | 53 | (11.2) |
| Organic disorders | 7 | (3.0) | 11 | (4.5) | 18 | (3.8) |
| *Other clinical characteristics* |  |  |  |  |  |  |
| Duration of illness episode before admission <= 1 month, n (%) | 89 | (36.4) | 84 | (34.4) | 173 | (36.3) |
| Somatic comorbidity n (%) | 66 | (28.4) | 55 | (22.6) | 121 | (25.4) |
| Involuntary admission, n (%) | 40 | (17.2) | 42 | (17.2) | 82 | (17.2) |
| Admission last 2 years >= 1, n (%) | 108 | (46.6) | 114 | (46.7) | 222 | (46.6) |
| Previous criminal conviction, n (%) | 18 | (7.8) | 18 | (7.4) | 36 | (7.6) |
| Violence towards others past year, n (%) | 15 | (6.5) | 19 | (7.8) | 34 | (7.2) |
| Suicidality (>=1 previous suicide attempt), n (%) | 74 | (31.9) | 62 | (25.4) | 136 | (28.6) |
| Baseline CGI-S, median, (IQR) | 5 | (4 to 5) |  |  | 5 | (4 to 5) |
| Sleep-wake disruption n, (%)* | 112 | (48.3) | 5 | (4 to 5) | 209 | (44.0) |

Notes. Percentages are reported for all individuals regardless of missingness which ranged from 0 % for many demographic variables to 41 % for previous suicide attempt (detailed baseline characteristic tables found in Supplementary data) \*Sleep-wake disruption = Difficulties falling asleep, nocturnal awakenings, or early morning awakenings; 3 or more times per week.

#### Primary outcome

The primary outcome was mean duration of admission in days per individual. Admission was defined as the time and date of the initiation of the intake assessment. Time of discharge was defined as midday of the day the patient left the light environment to which they were randomized for > 24h. Duration of admission was calculated as the date and time of discharge minus the date and time of admission.

#### Secondary outcomes

Clinical outcomes

a) Clinical Global Impression (CGI) The CGI is a well-established, practical measurement tool that is widely used in RCTs, including studies of inpatient and trans-diagnostic samples.^27–29^ The CGI has two components, the improvement and severity scales, which together are used to quantify and track clinical progress and outcome.^30^ Clinical Global Impression, Improvement subscale (CGI-I): We used the improved version of the CGI-I.^31^ Scores can range from –6 (maximum deterioration) to +6 (ideal improvement), with a higher CGI-I rating indicating greater improvement relative to baseline status. The CGI-I was rated at discharge. A change of 4 or more on the CGI-I denotes a considerable improvement that is clear and clinically meaningful.^31^ Clinical Global Impression, Illness Severity scale (CGI-S): The CGI-S is rated on a 1-7 Likert with high scores indicating worse clinical status and/or functioning.^30^ The instruction states: Considering your total clinical experience with this particular population, how mentally ill is the patient at this time? (1=Normal, not at all ill; 7=Among the most extremely ill patients). A CGI-S rating of 3 or less denotes that the individual is mildly unwell or better relative to other acutely admitted patients.^30^ The CGI-S was rated at admission and discharge.
b) Risk of harm to self or others Brøset Violence Checklist (BVC): Aggressive behaviour was assessed three times per 24 hours using the 6-item BVC assessing the presence of six specific behaviours (e.g., irritability, physically and verbally threatening behaviour, etc.) on a binary scale (0= not observed, 1=observed). We analysed BVC sum scores >= 2 which has been shown to be a severity of aggressive behaviour which predicts short-term risk of inpatient violence.^32,33^ Staff Observation Aggression Scale-Revised (SOAS-R): Incidents of actual violence were systematically recorded using the SOAS-R^34^ after an incident had occurred.The SOAS-R total score ranges from 0 to 22 and a score >= 9 indicates that more serious incidents have occurred (e.g., inflicting physical pain or injury). Suicide Risk: suicidality was assessed and recorded daily.
c) Change in admission status. For patients who were involuntarily admitted, we assessed the time from intake to when admission status was changed from involuntary to voluntary admission.
d) Side effects The frequency and severity of side effects was rated on a 4-point scale using eight items measuring side effects of acute psychiatric treatments (e.g. difficulties with concentration, change in appetite) supplemented by the Headache and Eyestrain Scale (e.g. headache, dry eyes).^35^
e) Self-reported satisfaction Individuals routinely rate their satisfaction with an admission using a standard 11-item questionnaire (each item is rated from 1-5, with higher scores indicating greater satisfaction).

## Statistical methods

The analyses reported in the main text refer to the intent to treat (ITT) population.

## Sample size calculations

The sample size calculation is described in detail in the published protocol.^1^ Briefly, the calculation was performed for the comparison of the primary outcome, mean number of days hospitalized per individual exposed to the experimental or control lighting conditions. Assuming the experimental lighting conditions led to a reduction in the mean length of stay from about 6 to 5 days (with a standard deviation of about 3.5 days), then 194 participants in each condition would give an 80% chance at an alpha = 0.05 to detect a difference in the length of stay of one day using an ITT analysis.

## Statistical analysis

A detailed statistical analysis plan (SAP) was written and published prior to the unblinding of the dataset, and the data were analyzed according to this plan by statisticians prior to unblinding the dataset (see Supplementary Information, p. 40). Multivariable linear regression was used to analyze the effect of blue-depleted evening lighting on the duration of admission in days (primary outcome). These analyses were adjusted for a pre-specified set of baseline variables considered to be predictors of length of admission, including age, sex, psychiatric diagnosis, comorbid personality disorder or substance use disorder, whether the admission was voluntary or not, and the number and duration of previous admissions in the past 2 years. For the main analysis, the psychiatric diagnosis was categorized as either psychotic episode / disorder, manic episode, severe depression or other (see SAP for further details). As expected, duration of admission was right-skewed, and we used bootstrapping to obtain the 95 % confidence interval (CI) with the bias-corrected and accelerated (BC_a_) method and B=5000 bootstrap samples.

To assess if the effect of the blue-depleted evening lighting differed between subgroups, the regression analyses were repeated with an interaction term between each of the covariates listed above and randomisation group.

Secondary outcomes were analysed using linear regression models with the same covariates and bootstrapping approach as the primary variable: CGI-I and CGI-S at discharge, number of episodes with a BVC of 2 or more, number of episodes with a SOAR-S score of 8 or more, and side effects and satisfaction score. The remaining secondary outcomes were binary variables and were analysed using logistic regression with the same covariates as above. For change from involuntary to voluntary admission we additionally compared the lighting groups using a Cox regression model. The average score for each of the side effects scales are compared using two sample t-test, excluding individuals missing more than two items.

Post-hoc analysis: We estimated the Number Needed to Treat (NNT) to enable the investigators to interpret the clinical meaningfulness of any statistically significant between-group differences in key outcome measures (see Supplementary information, p. 64 for details of how it was calculated). We chose the NNT because it is widely used across medicine to communicate the effectiveness of different health care interventions and represents the average number of patients who need to be treated to prevent one additional bad outcome.^36^ In this RCT it is the number of patients that need to be exposed to the experimental lighting condition for one patient to benefit compared with the control condition. The user written package bcii in Stata/MP 18.0 was used to calculate the NNT with 95 % confidence intervals. This package employs the confidence interval calculation methods described by Bender (2001).^37^

### Study monitoring

A Data and Safety Monitoring Committee (DSMC) scrutinized trial progression, technical issues, and the safety of patients in weekly meetings and the DSMC offered guidance on the resolution of three issues (Supplementary information, p. 66).

### Funding

The study was funded by St. Olavs Hospital, NTNU, and the Council for Mental Health, ExtraStiftelsen.

## Results

Fig 1 provides an overview of the flow of participants through the clinical trial. The core characteristics of the intent to treat (ITT) sample and key clinical outcomes are summarized in Tables 1-2 and Fig 2-3 (see online supplementary information for additional details for all findings: Tables 2S-5S and Figure 2S).

**Fig. 2:**
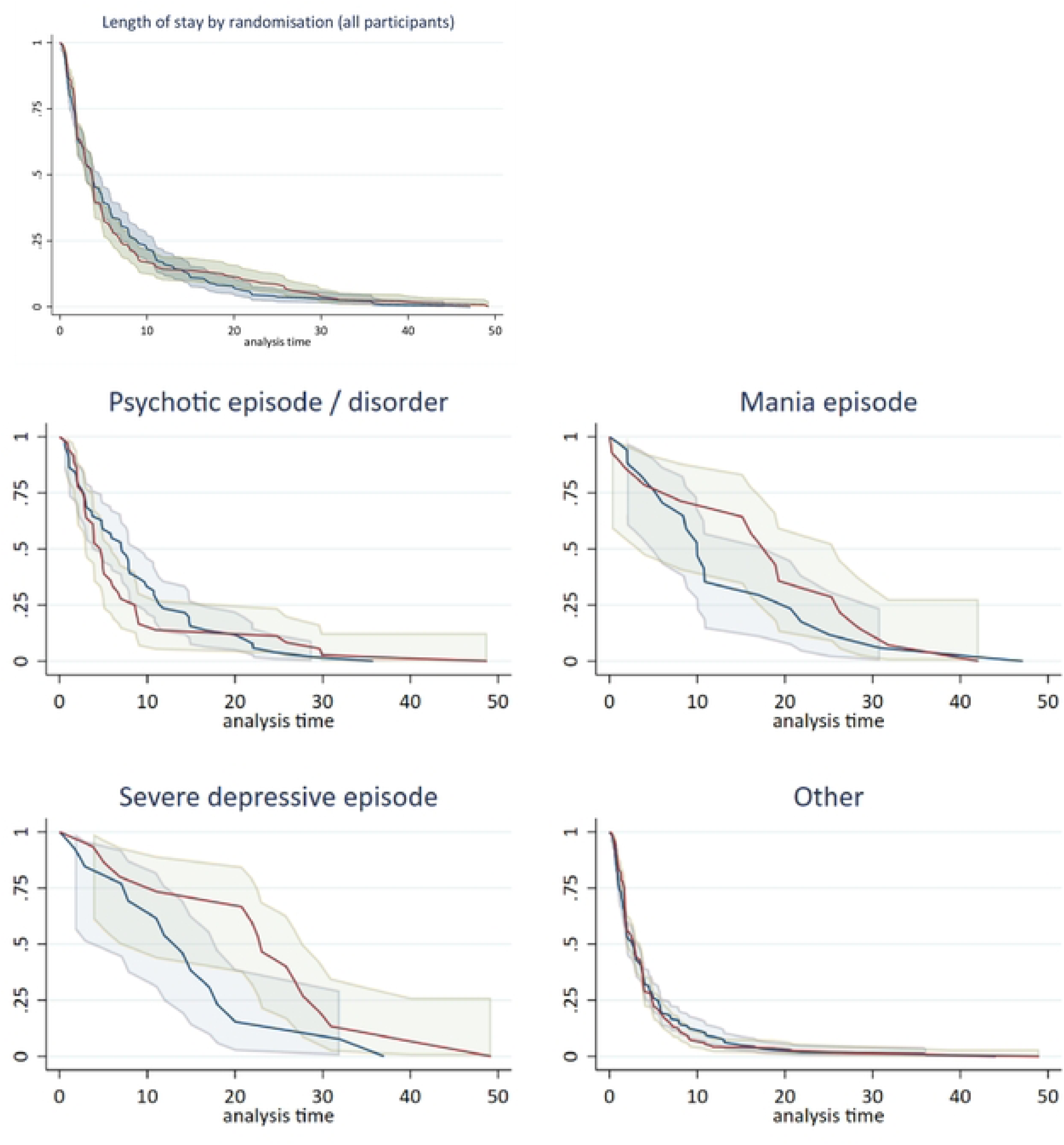
The proportion of patients who are still admitted by randomisation for pre-selected diagnostic categories. Red line= Evening blue depleted light environment. Blue line= Standard light environment. Panel A shows data for all participants <u>(N=476).</u> Panel B shows data for patients diagnosed with a psychotic episode (n=87). Panel C shows data for patients diagnosed with a manic episode (n=3I). Panel *D* shows patients diagnosed with a severe depressive episode (n=28). Panel E shows patients with all other diagnoses (n=329). There were no statistically significant differences between the subgroups (p=0.22).

**Fig 3.**
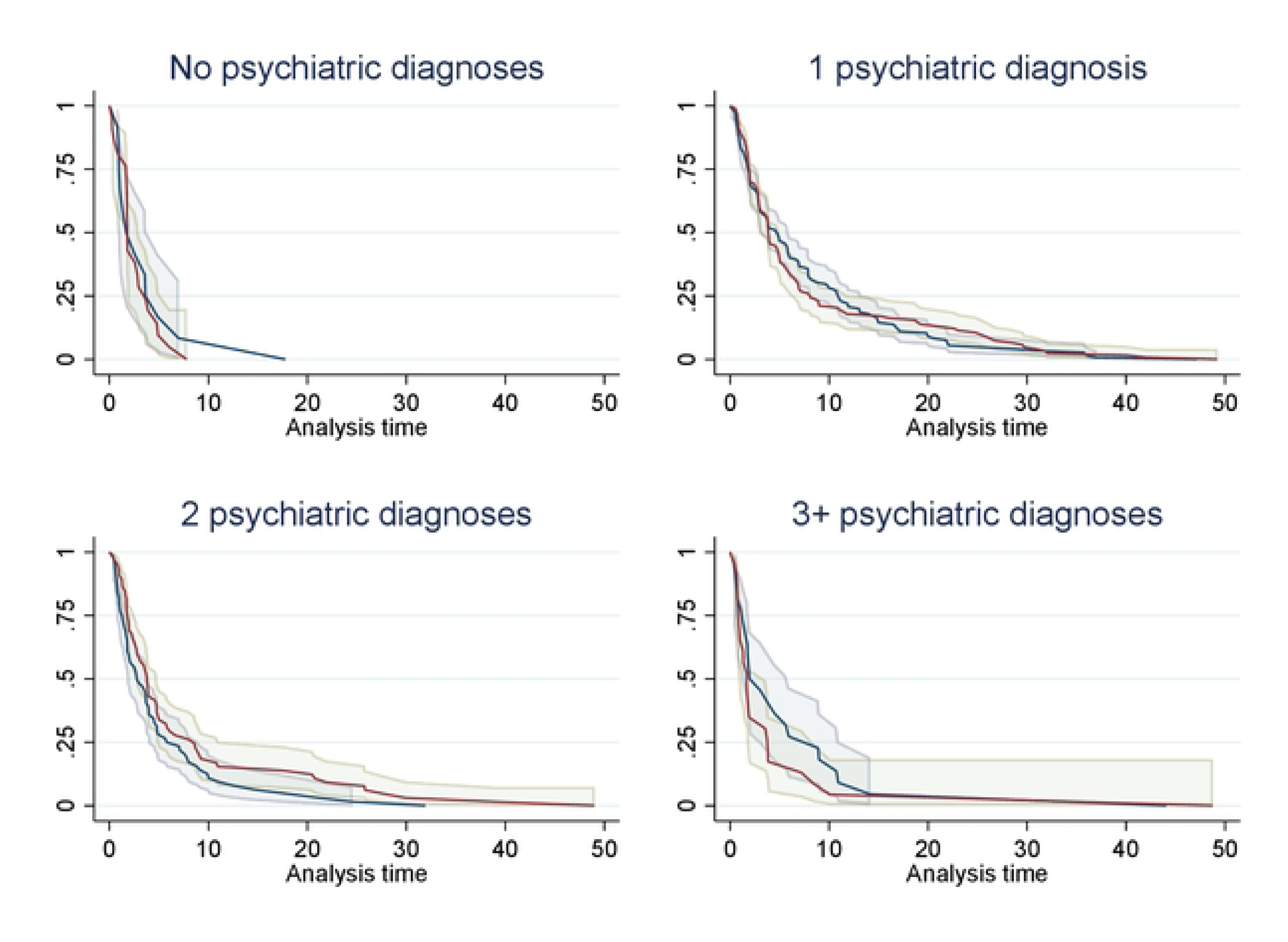
The proportion of all patients who were still admitted by randomisation for number of psychiatric diagnoses. Red line= Evening blue depicted light environment. Blue line= Standard light environment. Panel A shows data for no psychiatric diagnoses (n=33). Panel B shows data for patients with one psychiatric diagnosis (n=268). Panel C shows data for patients with two psychiatric diagnoses (n=129). Panel D shows patients diagnosed with three or more psychiatric diagnoses <u>(n=45).</u> There were no statistically significant differences between the subgroups (p=0.21).

**Table 2.** Primary and secondary outcomes for patients (N=476) in the Intention to Treat population, allocated to admission in a standard light environment (n=244) or evening blue-depleted light environment (n=232).

|  |  | **Blue-depleted evening light environment** | **Standard light environment** |  |  |
| --- | --- | --- | --- | --- | --- |
| **Linear regression** | **N** | **Estimated Mean (95% CI)** | **Estimated Mean (95% CI)** | **Mean difference (95 % CI)** | **p-value** |
| **Length of stay (days)** | **476** | **7.1 (6.1 to 8.1)** | **6.7 (5.8 to 7.5)** | **0.4 (-0.9 to 1.8)** | **0.523** |
| **Clinical Improvement during admission (CGI-I)** | 442 | 2.13 (1.94 to 2.31) | 1.85 (1.67 to 2.03) | 0.28 (0.02 to 0.54) | 0.035 |
| **Clinical state at discharge (CGI-S)** | 443 | 3.37 (3.26 to 3.48) | 3.55 (3.43 to 3.67) | -0.18 (-0.34 to -0.02) | 0.029 |
| **Aggressive behaviour (BVC)** |  |  |  |  |  |
| No. of serious events | 475 | 0.04 (-0.01 to 0.09) | 0.29 (0.17 to 0.41) | -0.25 (-0.38 to -0.12) | <0.001 |
| No. of serious events / 100 days | 475 | 0.31 (-0.08 to 0.71) | 3.30 (1.34 to 5.27) | -2.99 (-5.00 to -0.98) | 0.004 |
| **Violent Incidents (SOAS-R)** |  |  |  |  |  |
| No. of serious events | 476 | 0.18 (0.04 to 0.32) | 0.12 (0.05 to 0.18) | 0.07 (-0.09 to 0.23) | 0.414 |
| No. of serious events / 100 days | 476 | 0.97 (0.19 to 1.75) | 1.79 (0.34 to 3.24) | -0.82 (-2.55 to 0.91) | 0.353 |
| **Side effects** |  |  |  |  |  |
| Headache & eye strain scale | 215 | 1.99 (1.87 to 2.11) | 1.94 (1.82 to 2.06) | -0.04 (-0.21 to 0.12) | 0.599 |
| Other side effects | 217 | 2.00 (1.90 to 2.09) | 2.10 (1.99 to 2.18) | 0.09 (-0.05 to 0.22) | 0.199 |
| Total side effects | 217 | 1.99 (1.89 to 2.09) | 2.03 (1.93 to 2.13) | 0.04 (-0.09 to 0.18) | 0.532 |
| **Patient satisfaction<sup>a</sup>** | 205 | 3.77 (3.62 to 3.91) | 3.66 (3.52 to 3.80) | 0.10 (-0.10 to 0.31) | 0.322 |
| **Logistic regression** | **N** | **Estimated percentage (95% CI)** | **Estimated percentage (95% CI)** | **aOR (95 % CI)** | **p-value** |
| **Risk of suicide** | 412 | 42.2 (36.1 to 48.3) | 40.8 (34.8 to 46.8) | 1.07 (0.69 to 1.67) | 0.752 |
| **Required supervision due to risk of suicide** | 412 | 17.8 (13.0 to 22.7) | 20.1 (14.9 to 25.2) | 0.85 (0.49 to 1.44) | 0.538 |
| **Change from involuntary to voluntary status** | 79<sup>b</sup> | 27.5 (15.8 to 43.4) | 47.6 (33.0 to 62.7) | 0.43 (0.13 to 1.42) | 0.167 |
|  |  | **Predicted hazard ratio** | **Predicted hazard ratio** | **aHR (95 % CI)** | **p-value** |
| **Time to change from involuntary to voluntary status** | 82<sup>b</sup> | 0.20 (-0.11 to 0.50) | 0.27 (-0.15 to 0.69) | 0.73 (0.30 to 1.84) | 0.513 |
| *Notes.* iCGI-I: Clinical Global Impression Scale - Improvement (clinicians' assessment of improvement at time of discharge); CGI-S: Clinical Global Impression - Severity sub-scale (Severity at discharge); BVC: Brøset Violence Checklist (score $\geq 2$ considered "severe"); SOAS-R: Staff Observation Aggression Scale-Revised (score $\geq 9$ considered severe). <sup>a</sup>Ten-item satisfaction score with each item score from 1 to 6, averaged across all items with a response so long as 9 of 10 items are answered; <sup>b</sup>The adjusted logistic and Cox regression analyses includes only those who were admitted involuntarily at the review on day 2, additionally all three participants with admitted involuntarily with depression episode subsequently ended up with a voluntary admission status and were excluded from the logistic regression. | | | | | |

### Sample Characteristics

As shown in Table 1, the ITT sample comprised 476 individuals (Blue-depleted LE=232; Standard LE=244). The sample mean (SD) age was 37.2 (13.9) years, 139 were male (41%). 369 (78%) were of European origin, and 91 (19%) were full-time employed. The most frequent primary diagnoses were: affective disorder (n=121; 25%), 31 of which were in a manic episode and 28 had a severe depressive episode and psychotic episodes (n=87; 18%). Thirty seven percent of the sample (n=174) received two or more ICD-10 diagnoses. At admission, the sample mean CGI-S rating was 4.6 (median 5) and 174 (37%) had been unwell for >=30 days. See supplementary table 2S for details.

There were 121 (25%) individuals with at least one somatic condition and 209 (44%) reported current sleep disturbances. Notably, there was no record that 299 (63%) of the sample were receiving psychotropic medications at the time of admission. Of those receiving medications, the most frequently used medications were: antipsychotics (n=129, 27%), benzodiazepines/z-hypnotics (n=59, 12%), and anti-depressants (n=56, 12%).

### Clinical Outcomes

As summarized in Table 2, there were no statistically significant differences in the duration of hospitalization in individuals allocated to the evening Blue-depleted LE (Mean: 7.06; Median: 3.71) as compared with the Standard LE (Mean: 6.72; Median: 3.68), nor duration of hospitalization according to diagnostic subtypes (p=0.22) or number of diagnoses per individual (p=0.21) (Fig. 2 and fig 3). See supplementary information, table 3S for observed values, and table 4S for subgroup analyses for duration of hospitalization. There were no relevant differences in duration of hospitalization between the ITT and PP samples (see Supplementary information, table S5).

Compared with the Standard LE, individuals allocated to the Blue-depleted LE showed significantly higher improvement during admission (Clinical Global Impressions scale-Improvement, CGI-I, estimated mean difference: 0.28, 95% CI: 0.02,0.54; p=0.035), Number Needed to Treat (NNT): 12 (95 % CI 6 to 61); and lower illness severity at discharge (Clinical Global Impressions Scale-Severity of illness, CGI-S, estimated mean difference: –0.18, 95% CI: –0.34 to –0.02; p=0.029, NNT: 7 (95 % CI 4 to 22).

The Broset Violence Checklist (BVC) ratings of aggressive behaviours were significantly lower in the Blue-depleted LE compared with the Standard LE. The estimated difference in predicted serious events per 100 days: –2.98; 95% CI: –4.98 to –0.99; p=0.003, NNT: 9 (95% CI: 7 to 15).

There were no significant between-group differences in actual violence episodes, suicide attempts, or the probability of changing from involuntary to voluntary status. Likewise, side effects (see also supplementary fig 2S for details) and satisfaction ratings did not differ between groups (table 2).

## DISCUSSION

In this randomised controlled trial, we tested if there are any benefits of modifying the evening light environment (LE) in an acute psychiatric hospital according to chronobiological principles. We found no differences between the Blue-depleted LE and Standard LE in our primary outcome of duration of hospitalization. However, there were benefits observed by clinicians on ratings of clinical improvement (CGI) and aggressive behaviour (BVC). In terms of clinical significance, the Numbers Needed to Treat (NNTs) for greater clinical improvements and lower levels of aggressive behaviour are similar to other low intensity interventions employed in psychiatry and in general medicine that are included in NICE guidelines. Further, these clinical benefits were achieved without any discernible differences in patient-rated side effects or treatment satisfaction levels. Moreover, we did not find any association with lighting conditions and transition from involuntary to voluntary status, nor any differences in actual violent events or suicidality, though these were low-frequency events and it is likely that the trial was insufficiently powered to detect these events.

We acknowledge that our primary outcome measure and the design of this RCT was ambitious. Nevertheless, we believe these choices were justified as inpatient admissions are a burden to patients and the largest contributor to the cost of psychiatric care and a reduction in bed-days would not only be welcomed by patients but also offset the cost of new lighting systems, and we wanted to test if there were any benefits of the lighting conditions in a real-world clinical setting. A defining characteristic of a public health system that takes all admissions from a defined catchment area is that the inpatient population is heterogeneous, and the throughput of admissions is rapid, with short lengths of stay being the norm. This was true of our study setting with 1118 admissions over eight months and a median duration of admission of only about four days. Employing a pragmatic effectiveness design with deferred consent meant that this RCT recruited nearly half of all the patients admitted from a population of 300,000 people, including involuntary admissions and many other individuals with severe or complex problems who would normally be excluded from research studies. As such, this RCT has greater external validity, and findings are more generalizable, than efficacy studies undertaken in specialist clinics and other selective research settings that tend to recruit small homogenous samples.

Whilst we set a very high threshold for finding between-group differences in the duration of acute admissions, it is notable that this is the third RCT exploring lighting in psychiatric inpatient settings that has failed to find a significant reduction in the length of admission.^19,20^ Length of acute admission and decisions about hospital discharge are not only based on the clinician and patient observations regarding progress, but also on structural and practical issues, such as vacancies for suitable placements at other hospital facilities units and the need to accommodate new acute admissions. Whilst the duration of admission was an obvious choice as the primary outcome, it is also true that these other factors may have reduced the precision of the findings. The absence of significant between-group differences in outcomes related to involuntary status, suicidality or violent incidents may be explained as these were low-frequency events, which reduced the statistical power to find differences. As such, we can only conclude that exposure to the blue-depleted LE did not exacerbate any of these problems.

The key differences we identified between the light environments were that exposure to the blue-depleted LE was associated with better clinical evaluations of outcome, greater improvement, and lower levels of aggression during admission. The NNTs were between 7 and 12 for these outcomes, which matches that of other low-intensity interventions in clinical psychology or medicine.^38,39^ The CGI-S and CGI-I are widely employed in RCTs in inpatient units and have clearly defined anchor points,^27,28,30,31^ and the BVC is a measure of observed behaviour that has been validated and implemented in more than 20 countries.^40^ Our findings, therefore, suggest some cause for optimism that modifying the evening light environment in a psychiatric inpatient unit could benefit a wide range of acute admitted inpatients. However, we propose two potential research directions that may enhance the outcomes in the future: (i) determining the optimal intervention dosage and enhancing adherence, and (ii) balancing global versus specific outcome measures and beginning to disentangle moderators and mediators of response.

We have previously shown that the same light environment as employed in the current RCT produced changes in sleep, circadian rhythms, and arousal in healthy young adults.^17,18^ Further, there were no negative effects in that population, nor in nurses’ ability to work in the blue-depleted LE at the present unit,^41^ or elsewhere.^42^ However, it is possible that aspects of the ‘dosage’ we employed is suboptimal. The evening light in the current trial was between 7 and 21 melanopic lux,^18^ whereas new guidelines suggest that 10 melanopic lux is the maximum intensity to avoid negative impact on the circadian system in healthy individuals.^43^ The melanopic lux we employed is lower than in previous trials,^19,20^ but we were mindful that patients in an acute psychiatric ward may have an altered light sensitivity compared with the normal population.^44–47^ Moreover, we note that Henriksen et al. found a large effect on manic symptom severity after 7 days of evening use of blue-blocking glasses (BBG) as an adjunct to standard treatments, which is a longer exposure than most patients in our trial. It has also been argued that combining chronotherapeutic interventions may be synergistic,^21,48^ although our ambition was to test the isolated benefit of only modifying the evening light environment. It will be important for future research to test if changing the dose by increasing the number of days the patients are exposed and/or by adding other chronotherapeutic interventions, such as morning bright light exposure in non-manic patients, will influence key clinical outcomes.

On a related theme to dosage, is the issue of adherence with the intervention. Previous trials of psychiatric hospital lighting have been limited to single hospital rooms^20^ or have had limited control over other light sources^19^ that could impact adherence. We had a specific focus on securing adherence, such as installing the light environment in patient rooms, bathrooms, and common areas in unit, deploying filters in front of windows in patient rooms after 1810h and permanent filters on TV screens, and offering blue-blocking filters on mobile devices. Lights in the outside recreation/smoking area were blue-depleted and patients were offered BBG whenever they left the ward. However, patients may have chosen not to use the BBG, and other adherence issues could have attenuated the observed effects. It is currently unknown how long exposure to ordinary evening light is sufficient to disrupt the effect of evening blue-depletion.

Many RCTs of neuropsychiatric patients employ CGI ratings to measure outcomes, even in disorder-specific studies.^27,29,49,50^ The advantage is that it is a clinically valid measure that clinicians can incorporate into day-to-day practice with minimal disruption.^30^ These global outcome measures are especially useful in large-scale pragmatic RCTs with heterogenous samples. The 476 participants in the current trial reported 125 different ICD-10 primary and secondary diagnoses with many presenting with comorbid mental and physical disorders. It was unfeasible to employ disorder-specific rating scales for such a broad and complex range of clinical presentations. However, using global outcome measures to assess the benefits of relatively low-intensity interventions risks missing subtle but important effects the intervention may have on both specific symptoms and/or transdiagnostic processes such as sleep-wake regulation.^13^ Further studies should strive to determine how specific patient characteristics, including mental disorder type, nature of sleep-wake disturbances, illness episode duration, and the interaction with certain medications^44–47,51,52^ may influence the efficacy of the intervention. Additionally, age-related factors could also modulate the intervention’s effectiveness.^53^ This complex array of potential moderators underscores the challenges clinicians currently face in assessing patient suitability for this intervention and may ultimately influence whether the intervention is employed in general psychiatric settings or restricted to specialist units.

In addition to the above issues, other limitations should be acknowledged. First, the primary outcome was heavily skewed. As described in the pre-publication Statistical Analytical Plan, this was expected, and bootstrapping was used to address this issue. The nature of this study meant it was impossible to blind clinicians to the lighting condition, but although we cannot rule out a potential bias, we have no evidence that conscious or unconscious bias would explain these findings, either in the decision to discharge a patient or in any of the clinical evaluations. The use of deferred consent meant that some patients left the unit before they could be approached regarding study participation and others did not sign the consent form. Therefore, about 50% of all admitted patients were included in the ITT population. There was a variable degree of data incompleteness at baseline, which may be attributable to the patients’ compromised capacity to provide data.

## Conclusion

Changing the evening light environment moderately enhances the clinical outcomes of standard acute psychiatric hospital inpatient care and treatment with estimated NNTs between 7 and 12 for clinical state, improvement, and aggressive behaviours. These benefits, coupled with the absence of side effects and the low intensity of the intervention, indicate that there is a reasonable case for broader adoption of this strategy, particularly in new units where the lighting system is being installed for the first time and is not replacing previous systems. Of course, further research is required on the nature and magnitude of benefits of evening blue-depleted LE and future research should consider whether the dose of light exposure and adherence to the condition improves patient outcomes, reduces length of stay, and/or identifies specific patient groups or symptoms that benefit more than others from admission in a blue-depleted LE.

## Data Availability

Study participants did not consent to have their data shared publicly. Deidentified participant data can potentially be made available after publication of the final version of the trial to researchers from accredited research institutions. Access to data will be limited to investigators who provide a methodologically sound proposal and will be for a specified time period (commencing about 3 months after publication and ending after 3 years). To ensure GDPR compliance, data processing must be covered by the 'Standard Contractual Clauses' from the European Commission, that data requesters have to sign. Proposals and requests for data access should be directed to and must be approved by the Data Protection Officer at St. Olavs Hospital and the Regional Committee for Medical and Health Research Ethics in Central Norway before any data can be shared.

## Acknowledgements

The authors would like to thank the patient user group, Regionalt brukerutvalg Helse Midt-Norge, for its contributions when designing the study, and the staff at the acute psychiatric unit at St. Olavs Hospital for their contributions to the study.

